# Maternal mortality risk among Black women in Brazil: a nationwide spatial consensus approach

**DOI:** 10.64898/2026.10.31.26366981

**Authors:** Ligia Vizeu Barrozo, Luana Silva Rodrigues de Araujo, Edson Amaro

## Abstract

Although preventable, maternal mortality remains a global challenge. Its burden is disproportionately concentrated in low- and middle-income settings. Substantial subnational geographical variations also persist within countries, driven by underlying socioeconomic disparities and unequal access to quality healthcare services. Here we propose a spatial consensus framework combining frequentist and Bayesian models to overcome variance instability in small-area estimates, mapping maternal mortality risk among Black women in Brazil to guide equitable public health interventions.

Using nationwide surveillance data (2019–2024), we applied a spatial Poisson probability model to detect high-rate clusters of maternal mortality among Black women. Concurrently, Bayesian hierarchical spatial models were fitted to smooth small-area rates and stabilize variance. Results from both approaches were integrated into a five-level risk typology to categorize Brazilian municipalities.

National aggregate reductions in Brazilian maternal mortality mask profound, spatially concentrated racial inequities. The spatial classification of risk scenarios identified 13 municipalities with Confirmed Critical Risk, 28 municipalities with Persistent Regional Risk, 12 municipalities with a Local Excess Alert, and 4 exhibiting Epidemiological Silence. The vast majority remained at the Expected Risk Level.

The proposed spatial consensus framework translates complex spatio-temporal modeling into an operational typology for surveillance systems. This approach bridges the gap between spatial analytics and public health decision-making, providing a replicable tool to address structural racial disparities in maternal health within decentralized health systems.

**Funding:** Institutional Development Program of the Unified Health System (PROADI-SUS), under protocol number (NUP) 25000.082775/2024-00.

## Introduction

In 2020, an estimated 287,000 women died from maternal causes globally, averaging nearly 800 deaths per day.^1^ A maternal death refers to any mortality occurring during gestation or within 42 days post-pregnancy attributable to obstetric causes, excluding accidental or incidental occurrences.^2^ Maternal mortality is quantified by the Maternal Mortality Ratio (MMR), defined as the number of maternal deaths per 100,000 live births within a given time period. Because the vast majority of maternal deaths stem from preventable obstetric complications, a high MMR directly reflects systemic failures in healthcare infrastructure, timely access, and quality of emergency care.^1^ Although preventable, maternal mortality remains a global challenge. Its burden is disproportionately concentrated in low- and middle-income settings: in 2020, the maternal mortality ratio was estimated at 430 deaths per 100,000 live births in low-income countries, compared with 13 per 100,000 in high-income countries.^1^ Moreover, substantial subnational geographical variations also persist within countries, driven by underlying socioeconomic disparities and unequal access to quality healthcare services.^3–5^

In Brazil, up to 92% of these deaths are considered avoidable.^5^ Since the creation of the Unified Health System (SUS), Brazilian MMR declined from 143·2 in 1990 to 59·7 per 100,000 live births in 2015,^6^ eventually stabilizing at approximately 52 to 55 per 100,000 live births in recent post-pandemic figures.^7^ Despite this substantial three-decade decline, Brazil remains far from reaching the target established by PAHO/WHO of fewer than 30 maternal deaths per 100,000 live births by 2030.^8^

National aggregate estimates, however, obscure substantial structural disparities. The disproportionate burden of maternal mortality among Black women in Brazil should not be interpreted as evidence of intrinsic biological susceptibility; rather, it reflects differential exposure to racial inequalities, barriers to timely and high-quality obstetric care, and preventable direct obstetric complications,^9^ reflecting entrenched racial inequalities, barriers to timely and quality obstetric care, and heightened exposure to preventable direct complications. Consequently, achieving health equity extends beyond considerations of optional normative objective, but a practical prerequisite for improving health outcomes, health system efficiency and for national progress.

Routine public health surveillance typically relies on conventional frequentist indicators, such as raw Standardized Mortality Ratios (SMRs). However, in nationwide contexts like Brazil, severe demographic heterogeneity introduces extreme variance instability. Municipal volumes of live births range from single digits to hundreds of thousands, generating a Coefficient of Variation well above 500% around a heavily right-skewed distribution. Under such population asymmetry, a single maternal death in a small municipality can spuriously inflate raw rates, introducing substantial spatial noise into geographic prioritization. To mitigate this instability, Bayesian spatial smoothing techniques, such as Conditional Autoregressive (CAR) models, are widely employed to ’borrow strength’ across neighboring jurisdictions, effectively stabilizing variance while capturing broad spatial trends.^10,11^

Despite its advantages, Bayesian spatial smoothing exhibits inherent methodological limitations. By shrinking local rates toward neighborhood means, CAR models carry a risk of oversmoothing, which can blur spatial boundaries and mask localized, isolated high-risk spikes, potentially generating false negatives in small, high-vulnerability municipalities.^12^ Furthermore, while Bayesian disease mapping provides continuous risk estimates and posterior probability distributions, it does not perform formal spatial hypothesis testing to evaluate the statistical significance or discrete boundaries of specific geographic clusters.^13^

To overcome these limitations, frequentist spatial scan statistics offer a complementary framework focused on localized cluster detection. Rather than smoothing rate distributions, spatial scan statistics utilize rigorous hypothesis testing, such as likelihood ratio tests via Monte Carlo simulations, to identify discrete spatial clusters of elevated risk without altering underlying local variation. Although early spatial scan methods were constrained to circular windows,^14^ recent advances, including the elliptic^15^ and flexible scan statistics,^16^ effectively capture irregularly shaped spatial clusters, demonstrating superior performance in detecting rare health events across vast territories.^17^

Because neither framework alone fully addresses spatial risk without trade-offs, an integrative consensus framework is required. Here we propose a five-level risk assessment combining frequentist spatial scan statistics and Bayesian hierarchical smoothing to map maternal mortality risk among Black women in Brazil (2019–2024) and establishing consensus risk scenarios to guide equitable public health interventions.

## Methods

### Data

We used publicly available data of maternal deaths and live births by municipality occurred from 2019 to 2024. Maternal death data were retrieved from the SIM/DATASUS official maternal mortality dataset (DOMAT), which includes deaths after investigation by local Maternal Death Surveillance Committees. The utilization of this consolidated system is supported by national methodological benchmarks for maternal mortality monitoring in Brazil.^18^

Information on recorded race/color was obtained from the Mortality Information System (SIM) database, which categorizes this field into five official groups: White (*branca*), Black (*preta*), Asian (*amarela*), Mixed-race (*parda*), and Indigenous (*indígena*). Following official Brazilian demographic standards (IBGE) and the Statute of Racial Equality (Law No. 12,288/2010), the Black population (*população negra*) was operationalized as the combined aggregate of deaths among *preta* (Black) and *parda* (Brown/Mixed-race) women.^19^ This aggregation reflects their shared exposure to structural racism and related social disadvantages in Brazil, rather than implying ethnic or phenotypic homogeneity.

Data on live births used as denominators for calculating the MMR were extracted from the Live Births Information System (SINASC - *Sistema de Informações sobre Nascidos Vivos*). Denominators were disaggregated according to the mother’s self-reported race/color using the same official IBGE categories. The denominator for the combined Black population was operationalized as the combined aggregate of live births to mothers identified as *preta* (Black) and *parda* (Mixed-race). Both datasets from SIM and SINASC were downloaded using the R environment.^20^

To overcome rate instability associated with rare events and small population sizes at the municipal level, data were aggregated across the entire six-year study period (2019–2024). Multi-year aggregation is a standard epidemiological approach in spatial disease mapping that accumulates sufficient statistical power to stabilize risk estimates and reveal persistent, structural geographic patterns rather than short-term stochastic fluctuations.^11,21^ Furthermore, while annual temporal variations, such as the mortality peak observed during the COVID-19 pandemic, were described epidemiologically, spatial models were applied to the aggregated period to identify core areas of long-term vulnerability.

For municipal-level spatial analysis, 246 live birth records among combined Black mothers with state-aggregate municipal codes (ending in ’0000’, denoting unknown municipality of residence) were excluded due to the absence of corresponding geographic boundaries for mapping. These records were retained in national and state-level aggregate analyses to ensure accurate denominator estimation and avoid overestimating mortality rates.

### Analysis

#### Statistical analysis

In order to evaluate overall racial disparities in maternal mortality, aggregate MMRs were calculated per 100,000 live births for Black and White women across the full study period (2019–2024). The overall rate difference between the two groups was evaluated using an exact Poisson rate ratio test, estimating the Rate Ratio with its corresponding 95% confidence interval (95% CI) and two-sided p-value (*p*<0·05). Temporal dynamics within the primary population of interest was assesses by an annual trend analysis conducted for combined Black women calculating single-year MMRs from 2019 through 2024. This enabled tracking the trajectory of maternal mortality among Black women across the pre-pandemic baseline (2019), the pandemic surge (2020–2021), and the post-pandemic period (2024).

#### Spatial analysis

### Spatial scan statistics

As maternal mortality can be considered a rare type of mortality, the flexible scan statistic was used to identify clusters using the ‘rflexscan’ package^22^ in the R environment.^20^ Prior to spatial analysis, municipal polygon boundaries originally in the SIRGAS 2000 geographic coordinate system (EPSG:4674) were projected into a planar metric coordinate system (SIRGAS 2000 / Brazil Polyconic, EPSG:5880) to derive accurate municipal centroids and Euclidean distance matrices without spatial distortion. A spatial Poisson probability model was applied to search for high-rate clusters of combined Black maternal mortality using a first-order Queen contiguity criterion. The maximum cluster size was set to *K* = 14 contiguous municipalities, balancing computational efficiency across the national territory (*N* = 5,570) and sensitivity to localized diseases aggregation.^16^ Statistical significance was evaluated using 999 Monte Carlo replications (*p*<0·05), and secondary clusters were restricted to non-overlapping geographic boundaries.

### Bayesian spatial relative risk estimates

To stabilize the variance of small-area rates and map spatial risk variations, Bayesian hierarchical spatial models were fitted using the CARBayes package^23^ in R^20^.

#### Bayesian Spatial Model Specification

For each municipality *i* (*i* = 1, …, *N*, where *N* = 5,570), the observed count of maternal deaths among Black women, denoted by *Y_i_*, was modeled assuming a Poisson distribution:

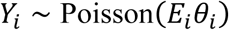

where *E_i_* is the expected number of maternal deaths under the nationwide baseline rate, and *θ_i_* represents the smoothed spatial Relative Risk (RR) for municipality *i*. The expected count *E_i_* was calculated as:

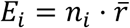

where *n_i_* is the total number of live births to combined Black mothers in municipality *i* over the study period, and 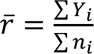 is the overall national maternal mortality rate for combined Black women.

#### Besag-York-Mollié (BYM) Spatial Prior Specification

The log relative risk log(*θ_i_*) was specified using the Besag-York-Mollié (BYM) model formulation,^10^ which decomposes spatial variability into a spatially structured random effect (*φ_i_*) and an unstructured random effect (*ψ_i_*):

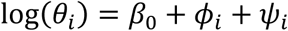

where:

- *β*_0_ is the overall intercept representing the overall log risk baseline.
- *ψ_i_* ∼ Normal(0, 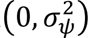) is an unstructured random effect capturing independent, area-specific random noise.
- *φ_i_* is a spatially structured random effect specified by an Intrinsic Conditional Autoregressive (ICAR) prior to model spatial dependencies across neighboring areas:

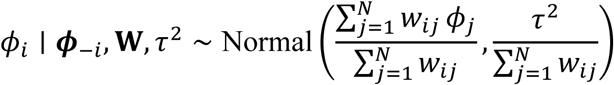

where **W** = (*w_ij_*) is an *N* × *N* spatial neighborhood matrix, defined using first-order queen contiguity (*w_ij_* = 1 if municipalities *i* and *j* share a common boundary or vertex, and *w_ij_* = 0 otherwise), and *τ*^2^ represents the variance parameter of the spatial random component.

#### Parameter Estimation and Inference

Model parameters were estimated using Markov Chain Monte Carlo (MCMC) simulation implemented via the S.CARbym() function in the CARBayes package.^23^ Posterior distributions were derived by running 120,000 iterations, discarding the initial 20,000 as burn-in, and retaining every 10th sample to minimize autocorrelation (thinning = 10). MCMC chain convergence for the baseline intercept was formally confirmed using Geweke’s diagnostic (*Z* = -1.49, p = 0.137), indicating that the post-burn-in iterations reached stationary equilibrium. Posterior mean estimates of *θ_i_* along with their 95% Credible Intervals (95% CrI) were calculated to construct smoothed relative risk maps across all Brazilian municipalities.

#### Epidemiologic scenarios

To synthesize spatial findings, municipalities were categorized into five risk scenarios, according to the combined results from scan statistic and Bayesian probability: (1) Confirmed Critical Risk, characterized by observed deaths (≥1) with consensus between high Bayesian probability (*P*[RR > 1] ≥ 0·95) and cluster assignment; (2) Persistent Regional Risk, with observed deaths (≥1) and high Bayesian probability (*P*[RR > 1] ≥ 0·95) without cluster assignment; (3) Local Excess Alert, with observed deaths (≥1) within a spatial cluster; and (4) Epidemiological Silence, defined as zero observed deaths despite high Bayesian posterior probability (*P*[RR > 1] ≥ 0·95) or spatial cluster membership, suggesting potential under-reporting; (5) Expected Risk Level, comprising all remaining municipalities without significant spatial excess.

#### Cartographic representation

All spatial representations were constructed in accordance with the semiology of graphics proposed by Bertin.^24^ Absolute counts of maternal deaths per municipality were represented using proportional circle maps, standardized rates and Bayesian estimates were mapped using choropleth maps, and cluster classifications and scenarios risks were depicted using chorochromatic maps.

### Ethics statement

Ethical approval was not required for this study, as it utilized secondary, anonymized, and publicly accessible aggregated data provided by the Brazilian Ministry of Health (DATASUS), in accordance with Resolution No. 510/2016 of the Brazilian National Health Council (CNS).^25^

## Results

From the initial 10,592 records extracted from the Mortality Information System (SIM), two cases were excluded because they did not occur within the pregnancy-postpartum cycle (TPMORTEOCO = 8), yielding a final analytical counts of 10,590 maternal deaths (overall MMR = 67.3 per 100,000), including 6,838 among combined Black women (MMR = 69.4 per 100,000 live births) and 3,337 among White women (MMR = 64.1 per 100,000 live births). A Poisson rate ratio comparison confirmed a statistically significant aggregate disparity (*p*<0.001), with combined Black women experiencing an 8% higher rate of maternal mortality overall compared to White women (Rate Ratio = 1.08, 95% CI: 1.04–1.13). However, multi-year aggregated rates were heavily inflated by the COVID-19 pandemic, during which both demographic groups experienced severe surges in maternal mortality, peaking in 2021 (111.5 per 100,000 for combined Black women and 121.3 per 100,000 for White women). As illustrated in Figure 1, combined Black women exhibited higher maternal mortality ratios across five of the six study years (2019–2020 and 2022–2024). Following the acute crisis of 2020–2021, rates fell sharply by 2022 and stabilized thereafter, reaching 55.9 per 100,000 live births in 2024.

**Figure 1.**
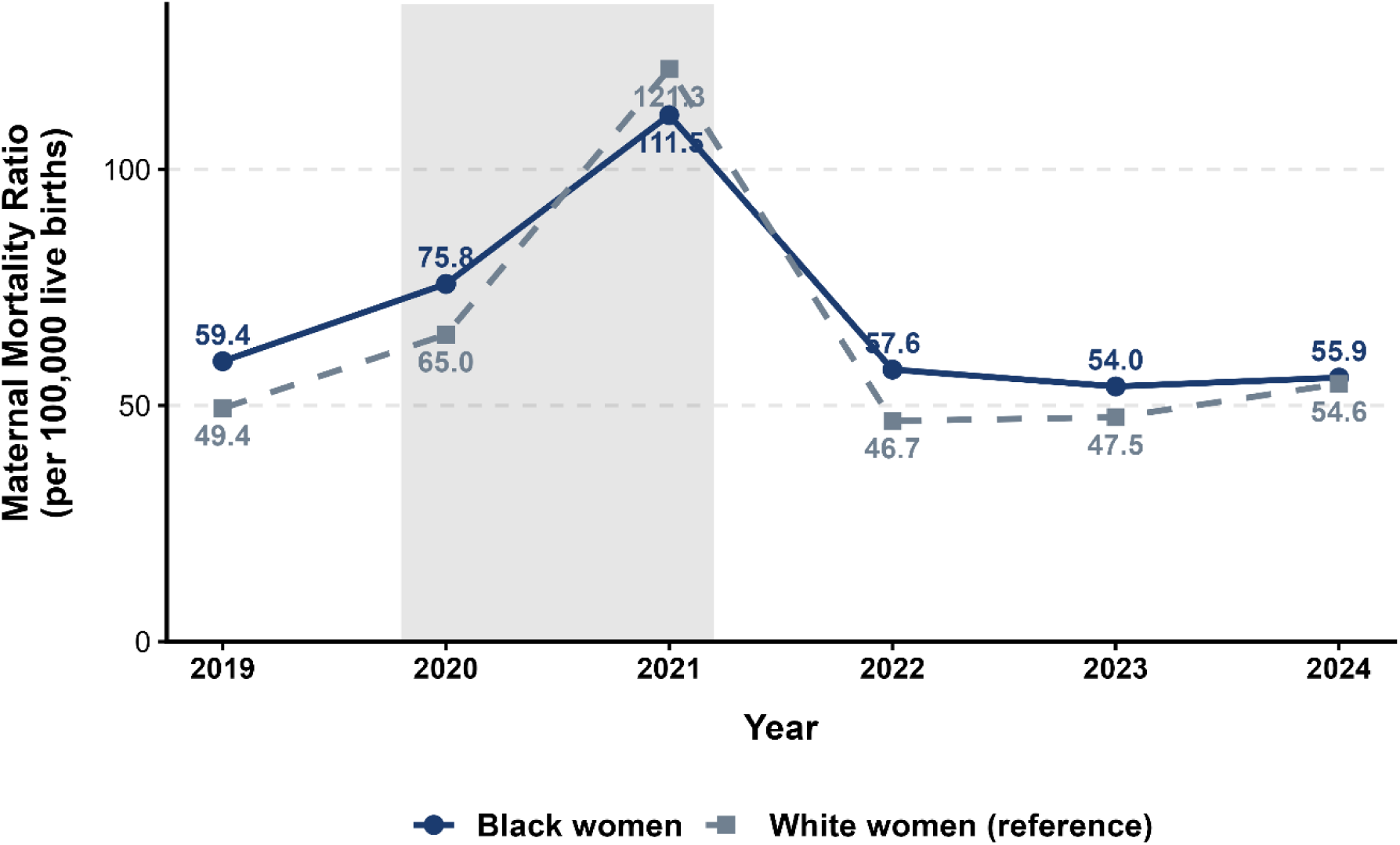
Temporal trends of the Maternal Mortality Ratio (MMR) among combined Black and White women in Brazil, 2019–2024. The shaded gray band highlights the peak impact period of the COVID-19 pandemic (2020–2021). Data sources: Mortality Information System (SIM/DATASUS) and Live Birth Information System (SINASC/DATASUS), Ministry of Health, Brazil.

Across the overall maternal deaths occurred from 2019 to 2024 (*N* = 10,590), the leading causes were indirect obstetric causes, followed by hypertensive disorders and obstetric hemorrhage. The proportional distribution of underlying causes between Black (*n* = 6,838) and White (*n* = 3,337) women differed significantly (ꭓ^2^ = 76.68, df = 7, *p* < 0.001). Although indirect causes predominated in both groups, they accounted for a substantially larger share of deaths among White women (45.3%, *n* = 1,513) than among Black women (37.9%, *n* = 2,589). Post-hoc analysis using adjusted standardized residuals demonstrated that Black women had significantly higher proportions of deaths driven by preventable direct obstetric conditions, specifically hypertensive disorders (*p* < 0.001), puerperal infection/sepsis (*p* = 0.001), and pregnancy with abortive outcome (*p* = 0.042). Conversely, White women presented a significantly higher proportion of deaths attributed to indirect obstetric causes (*p* < 0.001), whereas no statistically significant racial disparities in proportional distribution were observed for obstetric hemorrhage (*p* = 0.447), other direct complications (*p* = 0.555), or unspecified causes (*p* = 0.726).

To determine how these national disparities and preventable risks manifest geographically, this study first examined the unadjusted municipal counts and raw Standardized Mortality Ratios (SMR) among combined Black women. Municipal death counts ranged from 0 to 251 (highest in the municipality of Rio de Janeiro, State of Rio de Janeiro) (Fig. 2, left), while SMRs ranged from 0 to 90 (Fig. 2, right). The highest SMR was observed in Nova Esperança do Sul (State of Rio Grande do Sul), driven by 1 observed death against 0.011 expected deaths among only 16 live births over the 6-year period, illustrating the extreme variance instability inherent to small population sizes.

**Figure 2.**
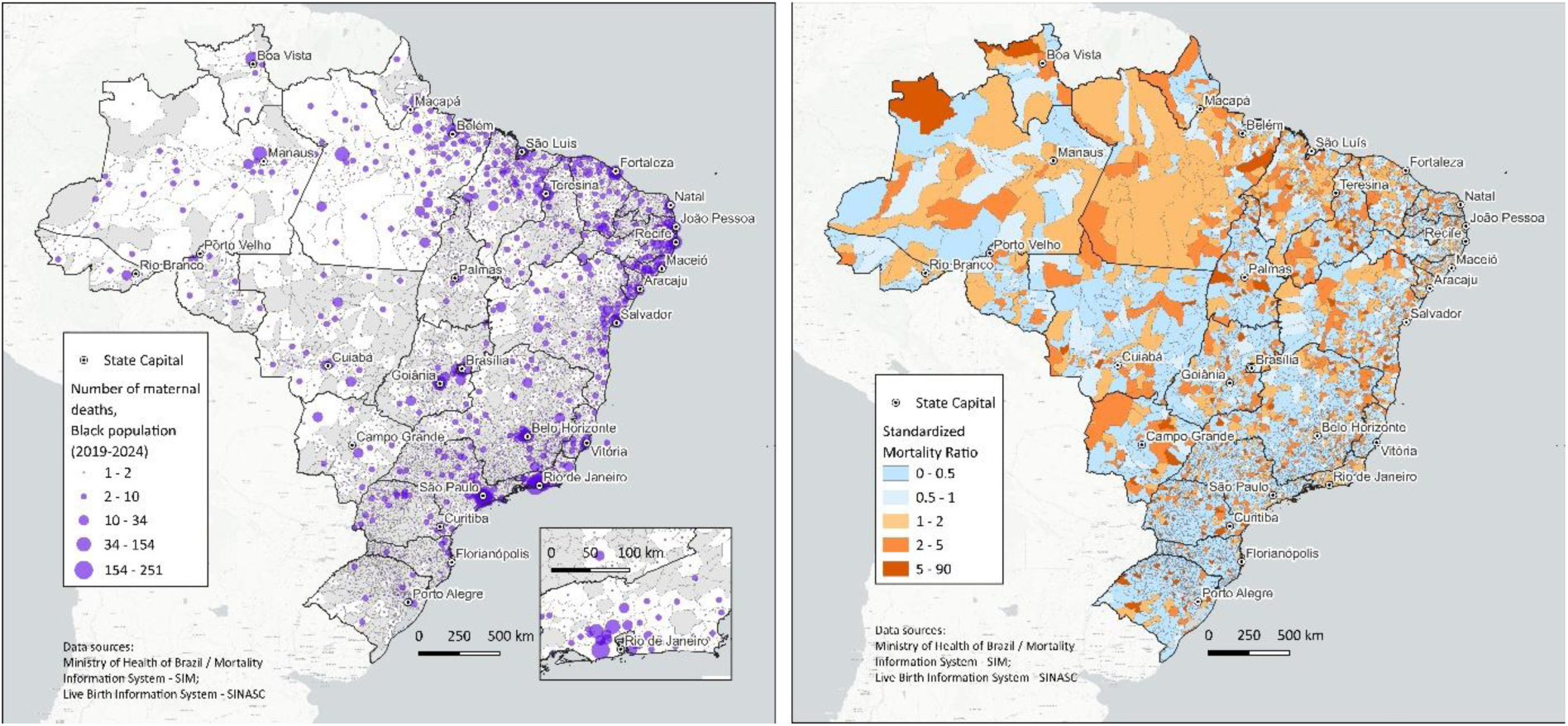
Spatial distribution of maternal deaths (left) and Standardized Mortality Ratio (right) among combined Black women in Brazil by municipality, 2019-2024. Data sources: DATASUS / Ministry of Health of Brazil. Geographic Coordinate System: SIRGAS 2000 (EPSG:4674).

Spatial smoothing via the Bayesian CAR model successfully eliminated stochastic noise and variance instability. Model convergence for the spatial Bayesian specification was verified via the Geweke diagnostic (Supplementary Figure S1). The distribution of z-scores remained centered around zero within ±1.96 threshold, confirming full convergence of the posterior distributions for municipal relative risk estimates. The posterior Bayesian spatial relative risk (RR) values ranged from 0.68 to 1.90 across municipalities (Figure 3, left).

**Figure 3.**
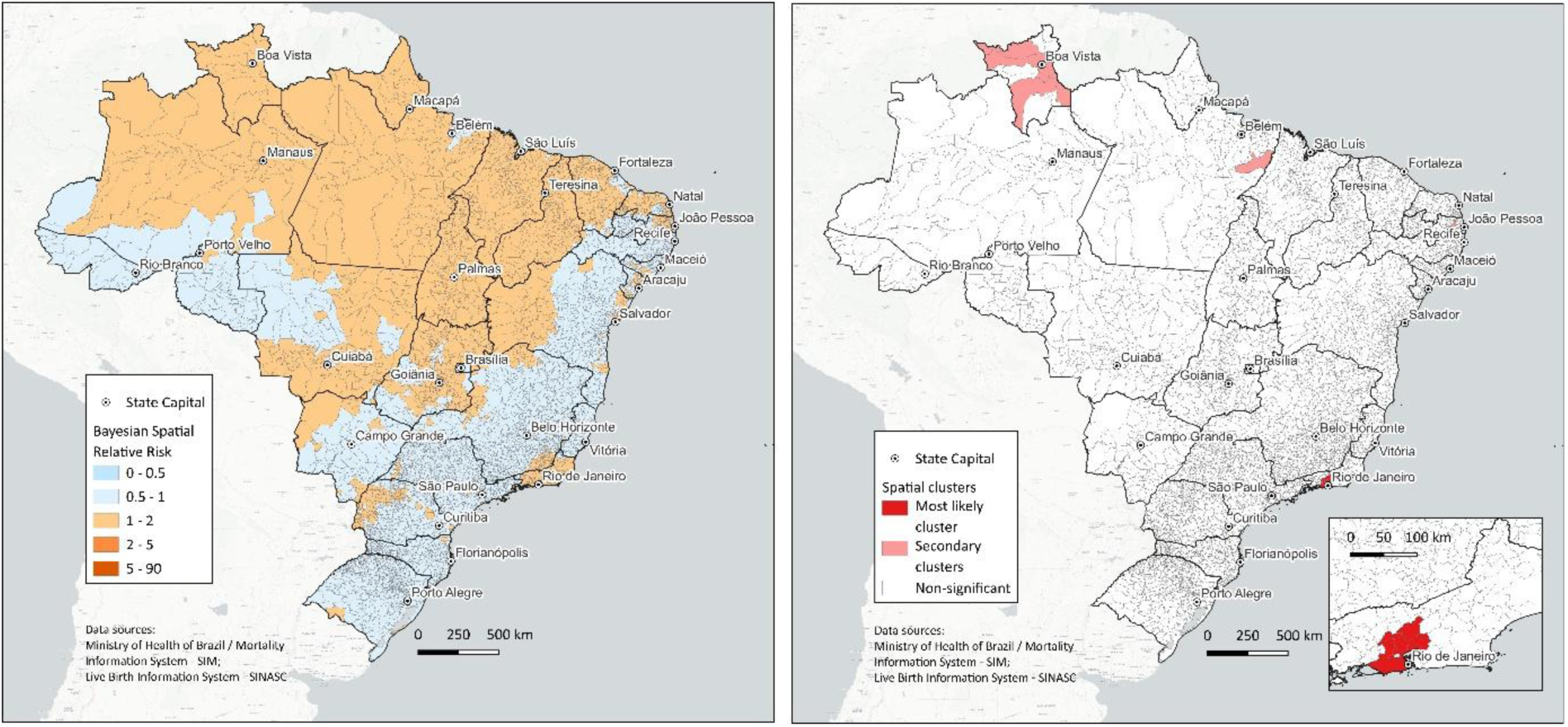
Bayesian spatial relative risk (left) and spatial clusters (right) of maternal mortality among combined Black women in Brazil by municipality, 2019–2024. Data sources: DATASUS / Ministry of Health of Brazil. Geographic Coordinate System: SIRGAS 2000 (EPSG:4674).

Spatial scan analysis using FleXScan identified three statistically significant spatial clusters of combined Black maternal mortality (*p*<0.05) (Figure 3, right). The primary (most likely) cluster comprised 10 contiguous municipalities in the State of Rio de Janeiro (*p* = 0.001), with 442 observed deaths compared to 295.8 expected deaths, representing a 49% excess risk relative to the national baseline (RR = 1.49). Two secondary clusters were also statistically significant: Cluster 2, consisting of 7 municipalities with more than double the expected risk (RR = 2.17, *O* = 68, *E* = 31.3, *p* = 0.004); and Cluster 3, a highly localized single-municipality cluster exhibiting an extreme relative risk (RR = 16.92, *O* = 7, *E* = 0.41, *p* = 0.042). The remaining 14 evaluated candidate clusters did not reach statistical significance (p ≥ 0.05).

In order to prioritize areas requiring urgent maternal health interventions we have classified the spatial patterns identified by model-based smoothing and spatial scan statistics into five consensus risk scenarios (Figure 4, Table 1). This integrative approach combines persistent smoothed relative risks (RR > 1.0) with local spatial clustering.

**Figure 4.**
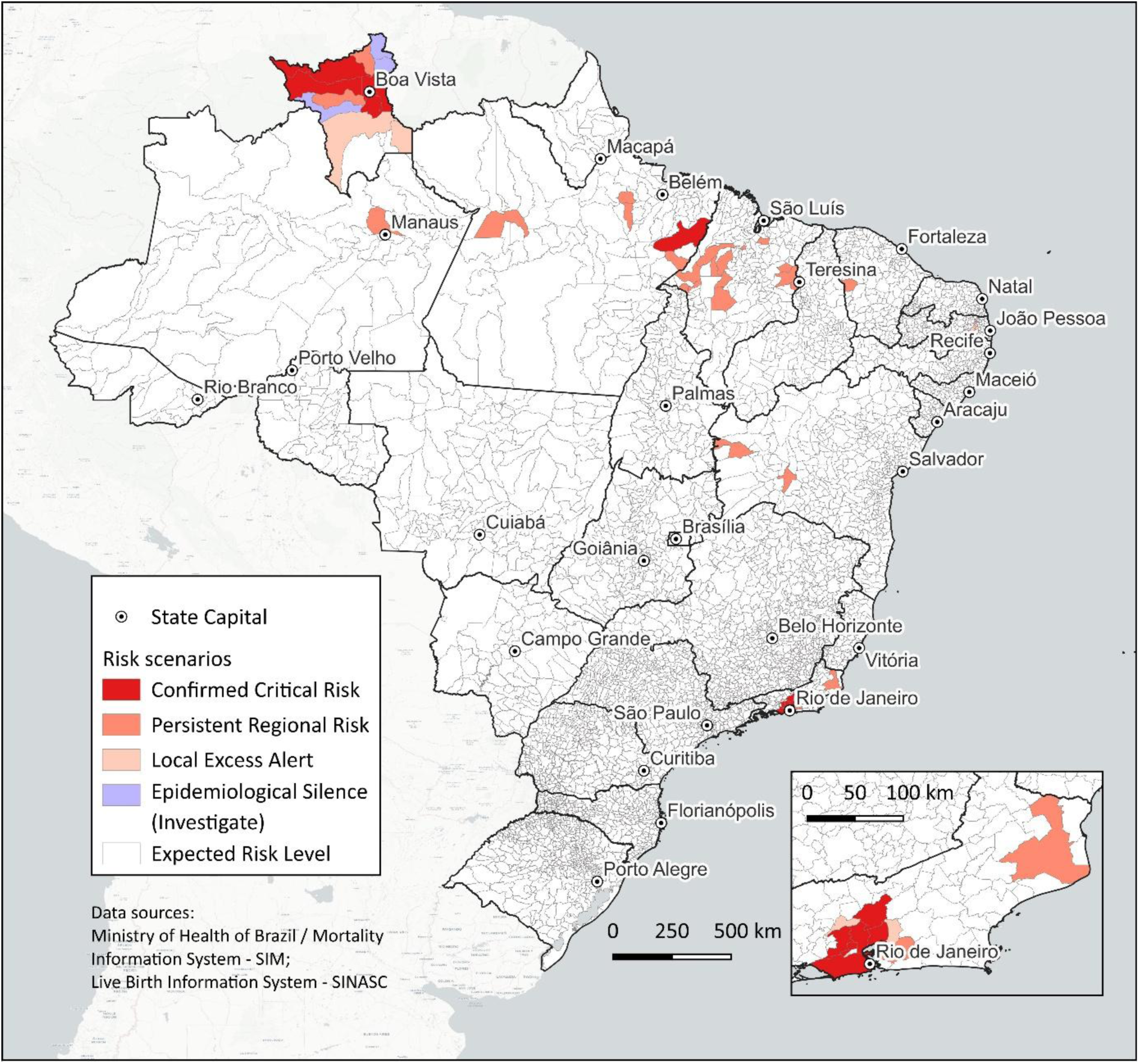
Spatial classification of risk scenarios for maternal mortality among combined Black women in Brazil by municipality, 2019–2024. Data sources: DATASUS / Ministry of Health of Brazil. Geographic Coordinate System: SIRGAS 2000 (EPSG:4674).

**Table 1.** Consensus risk scenarios for maternal mortality among combined Black women in Brazil (2019–2024): classification criteria, municipal distribution, and epidemiological indicators.

| Consensus Risk Scenario | Criteria Summary | Municipalities, *n* | Live Births, *n* | Observed Maternal Deaths, *n* | MMR |
|---|---|---|---|---|---|
| Confirmed Critical Risk | Deaths > 0 & posterior probability >= 0.95 & significant cluster | 13 | 459,254 | 504 | 109.7 |
| Persistent Regional Risk | Deaths > 0 & posterior probability >= 0.95 | 28 | 561,104 | 583 | 103.9 |
| Local Excess Alert | Deaths > 0 & significant cluster | 12 | 16,538 | 28 | 169.3 |
| Epidemiological Silence | Zero deaths & (posterior probability >= 0.95 OR significant cluster) | 4 | 1,253 | 0 | 0 |
| Expected Risk Level | Posterior probability < 0.95 & no spatial cluster detected | 5,513 | 8,813,973 | 5,723 | 64.9 |
| **Total** | **Brazil** | **5,570** | **9,852,122** | **6,838** | **69.4** |

The spatial classification of risk scenarios identified 13 municipalities with Confirmed Critical Risk, 28 municipalities with Persistent Regional Risk, 12 municipalities with a Local Excess Alert, and 4 exhibiting Epidemiological Silence (Table 1). The vast majority (*n* = 5,513) remained at the Expected Risk Level.

## Discussion

Maternal mortality in Brazil is still far from the target established by PAHO/WHO of fewer than 30 maternal deaths per 100,000 live births by 2030. Even in the end of the analysed period, the MMRs in 2024 reached 44.2 and 47.2 for White and combined Black women, respectively.

Counterfactual sensitivity analysis confirmed that the observed surge in the MMR among White women during the 2021 pandemic peak was substantially driven by a denominator artifact. Between 2019 and 2021, live births contracted by 9.90% among White women—a reduction 2.5 times sharper than that observed among combined Black women (-3.91%). This differential birth decline artificially added 10.32 deaths per 100,000 live births to the observed MMR of White women, compared to an effect of 3.56 points among combined Black women. When holding the live birth denominator constant at 2019 pre-pandemic levels, the counterfactual 2021 MMR for White women dropped from 104.3 to 94 per 100,000 live births, suggesting that differential denominator contraction contributed to the apparent racial inversion. Furthermore, while registered maternal deaths rose in both groups, the lower relative increase observed among combined Black women (+94·3% vs. +152·9% in White women) is consistent with potential ascertainment bias and differential underreporting during public health system collapse, as widely documented in the Brazilian context.^26,27^

Beyond spatial concentration, the stark divergence in the underlying causes of maternal death between racial groups exposes deep-seated inequities in the quality of clinical care. Over half of all maternal deaths among combined Black women (53.2%) stemmed from direct obstetric causes, conditions that are almost entirely preventable through timely, high-quality prenatal and emergency obstetric interventions. Conversely, indirect causes predominated among White women (45·4%), driven largely by the viral surge of COVID-19. This excess of direct obstetric mortality among combined Black women reflects systemic barriers in the continuum of care, including lower quality of prenatal coverage, delayed referral for high-risk pregnancies, and the structural phenomenon of obstetric ‘pilgrimage’, wherein marginalized women are forced to seek care across multiple hospital facilities during labor.^6,28^ Addressing racial disparities in Brazilian maternal mortality therefore requires not only geographic targeting, but also institutional reforms to eliminate racial bias in clinical decision-making and ensure equitable access to tertiary obstetric critical care.^29^

The substantial reduction in range from raw SMR to Bayesian spatial RR highlights the importance of spatial smoothing when analyzing rare health events in small populations. The remaining gradient (RR = 0.68 to 1.90) confirms that geographic disparities in combined Black maternal mortality are structural and spatially persistent, rather than artifacts of population size.

The integration of Bayesian spatial smoothing and flexible spatial scan statistics (FleXScan) into a unified classification matrix can, potentially, provide indications towards a robust decision-making framework for epidemiological surveillance and public health action. By cross-referencing model-based posterior probabilities with non-parametric cluster detection, municipalities were categorized into five operational risk scenarios, each reflecting distinct spatial dynamics, surveillance implications, and targeted intervention strategies. This approach allows a more direct basis to formulate context-based health action and/or policies within Brazilian healthcare system - SUS.

Methodologically, the convergence of smoothed model-based risks and non-parametric spatial scan statistics would confirm high spatial autocorrelation with maximum statistical certainty.^16,21^ From a public health perspective, these areas would face a severe, consolidated crisis in maternal care characterized by overlapping socio-spatial vulnerabilities and systemic healthcare failures. Surveillance would demand immediate, high-priority intervention, including reinforcing mandatory audits of 100% of maternal deaths by local committees, emergency resource allocation, and a comprehensive overhaul of local prenatal and emergency obstetric networks.^30^

The category of Persistent Regional Risk included 28 municipalities. Theoretically, the conditional autoregressive (CAR) framework captures broad, spatially structured trends that reflect diffuse regional risk factors, whereas spatial scan statistics may overlook non-compact or widely dispersed spatial patterns.^10,11^ For surveillance, this indicates a chronic, structural risk distributed across broader regional health jurisdictions, such as regional deficits in intensive care beds or defective emergency transfer protocols, rather than a localized outbreak. Recommended actions center on inter-municipal health governance, regional network reorganization, and strengthening regional referral pathways.

The scenario of Local Excess Alert comprised 12 municipalities. The FleXScan algorithm is uniquely sensitive to irregularly shaped, highly localized spatial aggregates that may not meet the threshold for long-term Bayesian spatial smoothing.^16^ Epidemiologically, this scenario flags acute, localized clusters or short-term spikes in maternal mortality, often tied to facility-level bottlenecks, localized clinical management errors, or temporary disruptions in local emergency services. Surveillance response requires rapid, local epidemiological investigations and facility-based audits to identify and rectify acute operational bottlenecks.

The category of Epidemiological Silence encompasses four municipalities. Theoretically, this scenario relies on the Bayesian CAR model’s ability to “borrow strength” from neighboring areas, revealing an underlying latent spatial risk that is masked by local zero counts.^10,11^ For epidemiological surveillance, this discrepancy signals a high probability of under-reporting, misclassification of maternal causes of death, or barriers to local healthcare access that force women to seek care in adjacent municipalities without spatial linkage to their true residence.^18^ Surveillance actions in these areas must prioritize active case finding of deaths among women of reproductive age, auditing hospital records, and training local vital registration teams.^30^

Finally, the Expected Risk Level category comprises all remaining municipalities where neither method detected significant spatial excess. Theoretically, these areas align with baseline national or regional expectations, representing stochastic variation without spatial aggregation.^11^ For surveillance, this class requires maintaining routine vital event monitoring, ensuring continuous reporting quality, and sustaining standard maternal health promotion policies.

Methodologically, this consensus framework bridges the gap between discrete cluster detection and continuous risk smoothing, filtering out variance instability while unmasking spatial vulnerabilities that a single method would miss. Operationally, it provides public health decisionmakers with a two-tiered strategy: immediate, targeted interventions and long-term structural planning.

This study has limitations inherent to spatial epidemiological designs utilizing secondary administrative data. First, despite using the consolidated DOMAT database and incorporating post-investigation reclassifications by Maternal Death Surveillance Committees, residual under-reporting and misclassification of maternal status on Death Certificates remain plausible, particularly in remote or under-resourced municipalities. Nevertheless, our framework explicitly addresses this constraint by establishing the ‘Epidemiological Silence’ scenario to systematically flag suspected under-reporting. Second, while race/color in SINASC is based on maternal self-report, race/color in SIM is typically proxy-reported by family members or hetero-identified by attending health professionals. This methodological discrepancy between birth and death registration systems may introduce racial misclassification bias or lead to missing data in secondary records.

This study demonstrates that national aggregate reductions in Brazilian maternal mortality mask profound, spatially concentrated racial inequities. Combined Black women consistently experienced higher MMRs than White women across the pre- and post-pandemic periods. Counterfactual analysis proved that the temporary 2021 spike observed among White women was an artifact of differential birth contraction rather than a true shift in racial vulnerability.

Achieving the Sustainable Development Goal target by 2030 remains unfeasible for Brazil without directly eliminating excess mortality among combined Black women. Public health authorities must leverage spatial consensus scenarios to prioritize emergency resource deployment, strengthen local Maternal Death Surveillance Committees, and enforce anti-racist, equitable clinical protocols across maternal healthcare networks.

## Supporting information

Supplemental Figure S1: Geweke convergence diagnostic

## Data Availability

All data used in this study are publicly available from the DATASUS portal (SIM and SINASC). The R scripts for spatial scan statistics and Bayesian hierarchical modelling are maintained in a dedicated repository and will be made publicly available on GitHub upon formal peer-reviewed publication. Scripts can be made available to researchers upon reasonable request.

## Contributors

LVB, EAJ and LSRA conceived the idea of the paper; LVB performed the statistical analysis; LVB, EAJ and LSRA wrote the preliminary version. All authors contributed to shaping and finalizing the manuscript. All authors approved the manuscript submission.

## Artificial Intelligence utilization

The author(s) acknowledge the use of an AI writing assistant for language refinement and editing support during the manuscript preparation. All data analysis, methodological implementation, interpretation of results, and final content validation were strictly conducted by the author(s).

## Data sharing statement

All data used in this study are publicly available from the DATASUS portal (SIM and SINASC). The R scripts for spatial scan statistics and Bayesian hierarchical modeling are maintained in a dedicated repository and will be made publicly available on GitHub upon formal peer-reviewed publication. Scripts can be made available to researchers upon reasonable request.

## Acknowledgement

The data or outcomes presented in this article were obtained through the project ‘VERACIS — Vulnerabilidades Étnico-Raciais, Ambiente, Clima e Impacto na Saúde’ (*VERACIS — Ethno-Racial Vulnerabilities, Environment, Climate, and Health Impact*), conducted by Hospital Israelita Albert Einstein in partnership with the Brazilian Ministry of Health, through the Institutional Development Program of the Unified Health System (PROADI-SUS), under protocol number (NUP) 25000.082775/2024-00.

