## Supplementary figures and images for "Maternal mortality risk among Black women in Brazil: a nationwide spatial consensus approach"

### Supplemental Figure S1: Geweke convergence diagnostic

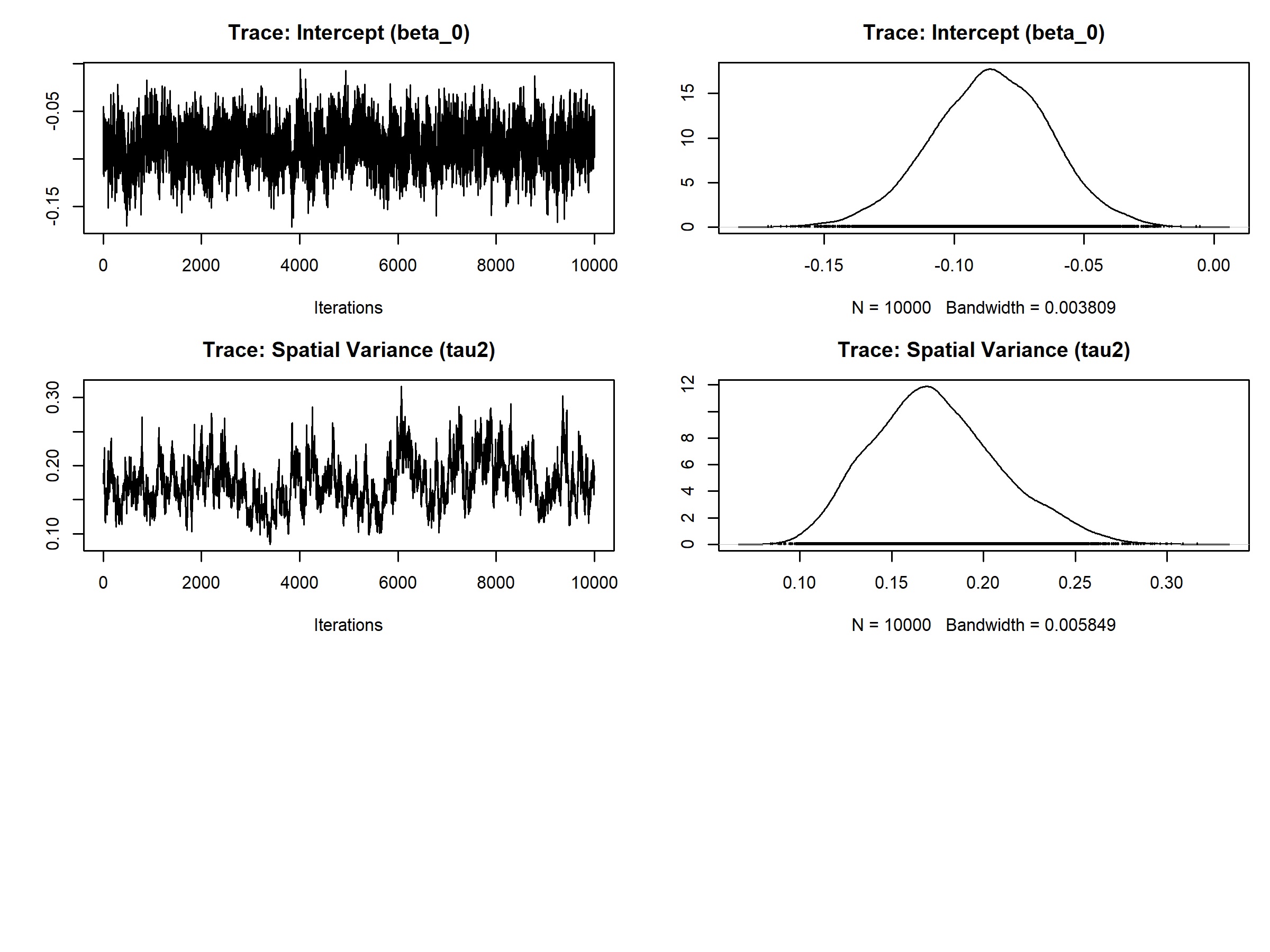
